## Supplemental Tables 1 and 2 for "Trends in social exposure to SARS-Cov-2 in France. Evidence from the national socio-epidemiological cohort – EPICOV"

### Supplementary Material

Table 1 Seroprevalence (ELISA-S $\geq$ 1.1[1]) according to *département* in November 2020 among people living in mainland France [2]. The national EpiCov cohort, 2020 November round

Table S2: Factors associated with seropositivity (ELISA-S $\geq$ 1.1[1]) in November 2020 among people living in mainland France [2]. The national EpiCov cohort, 2020 November round – Univariate and multivariate analysis including detailed occupation, detailed living conditions and self-reported distancing behaviours over the last 7 days

**Supplementary Table 1: Seroprevalence (ELISA-S ≥ 1.1[1]) according to *département* in November 2020 among people living in mainland France [2]. The national EpiCov cohort, 2020 November round**

| Department | Name Department | Region | N | n | % | 95% CI |
|---|---|---|---|---|---|---|
| 1 | Ain | Auvergne-Rhône-Alpes | 633 | 60 | 12.9% | [8.8-17.1] |
| 2 | Aisne | Hauts-de-France | 402 | 22 | 4.7% | [1.9-7.5] |
| 3 | Allier | Auvergne-Rhône-Alpes | 321 | 13 | 3.4% | [1.7-5.1] |
| 4 | Alpes-de-Haute-Provence | Provence-Alpes-Côte d'Azur | 286 | 13 | 3.0% | [1.3-4.7] |
| 5 | Hautes-Alpes | Provence-Alpes-Côte d'Azur | 325 | 20 | 4.7% | [1.8-7.6] |
| 6 | Alpes-Maritimes | Provence-Alpes-Côte d'Azur | 866 | 35 | 3.0% | [1.2-4.9] |
| 7 | Ardèche | Auvergne-Rhône-Alpes | 348 | 30 | 7.3% | [3.3-11.4] |
| 8 | Ardennes | Grand Est | 340 | 14 | 4.2% | [2.7-5.7] |
| 9 | Ariège | Occitanie | 276 | 7 | 1.6% | [0-3.9] |
| 10 | Aube | Grand Est | 287 | 15 | 5.8% | [3.4-8.2] |
| 11 | Aude | Occitanie | 313 | 14 | 3.8% | [1.4-6.1] |
| 12 | Aveyron | Occitanie | 305 | 8 | 3.5% | [0.6-6.3] |
| 13 | Bouches-du-Rhône | Provence-Alpes-Côte d'Azur | 1631 | 99 | 6.0% | [4.6-7.3] |
| 14 | Calvados | Normandie | 631 | 28 | 3.0% | [1.6-4.3] |
| 15 | Cantal | Auvergne-Rhône-Alpes | 293 | 9 | 2.4% | [0.8-3.9] |
| 16 | Charente | Nouvelle-Aquitaine | 343 | 7 | 2.5% | [1.1-3.8] |
| 17 | Charente-Maritime | Nouvelle-Aquitaine | 625 | 12 | 2.4% | [0.5-4.2] |
| 18 | Cher | Centre-Val de Loire | 312 | 15 | 5.2% | [2.9-7.5] |
| 19 | Corrèze | Nouvelle-Aquitaine | 297 | 13 | 5.1% | [2.4-7.8] |
| 21 | Côte-d'Or | Bourgogne-Franche-Comté | 529 | 31 | 5.8% | [3.5-8.1] |
| 22 | Côtes d'Armor | Bretagne | 539 | 16 | 1.7% | [0.8-2.7] |
| 23 | Creuse | Nouvelle-Aquitaine | 284 | 7 | 2.9% | [1.5-4.4] |
| 24 | Dordogne | Nouvelle-Aquitaine | 367 | 12 | 2.6% | [1.2-4] |
| 25 | Doubs | Bourgogne-Franche-Comté | 539 | 39 | 5.0% | [2.9-7.1] |
| 26 | Drôme | Auvergne-Rhône-Alpes | 486 | 32 | 6.9% | [4.1-9.7] |
| 27 | Eure | Normandie | 457 | 22 | 3.6% | [1.9-5.3] |
| 28 | Eure-et-Loir | Centre-Val de Loire | 357 | 18 | 5.6% | [2.8-8.5] |
| 29 | Finistère | Bretagne | 984 | 20 | 1.8% | [0.8-2.7] |
| 30 | Gard | Occitanie | 631 | 29 | 3.6% | [2.2-5] |
| 31 | Haute-Garonne | Occitanie | 1526 | 83 | 6.1% | [4.1-8.2] |
| 32 | Gers | Occitanie | 332 | 7 | 1.4% | [0.3-2.4] |
| 33 | Gironde | Nouvelle-Aquitaine | 1575 | 65 | 4.2% | [3.1-5.3] |
| 34 | Hérault | Occitanie | 1077 | 48 | 5.7% | [4.2-7.3] |
| 35 | Ille-et-Vilaine | Bretagne | 1159 | 38 | 2.7% | [1.6-3.8] |
| 36 | Indre | Centre-Val de Loire | 276 | 11 | 2.3% | [1-3.6] |
| 37 | Indre-et-Loire | Centre-Val de Loire | 604 | 18 | 2.4% | [1.4-3.4] |
| 38 | Isère | Auvergne-Rhône-Alpes | 1359 | 91 | 7.2% | [5.2-9.2] |
| 39 | Jura | Bourgogne-Franche-Comté | 324 | 19 | 7.3% | [3.9-10.7] |
| 40 | Landes | Nouvelle-Aquitaine | 377 | 15 | 4.9% | [2.9-7] |
| 41 | Loir-et-Cher | Centre-Val de Loire | 343 | 9 | 2.5% | [0-5] |
| 42 | Loire | Auvergne-Rhône-Alpes | 711 | 59 | 6.4% | [4.3-8.5] |
| 43 | Haute-Loire | Auvergne-Rhône-Alpes | 343 | 38 | 10.5% | [5.6-15.4] |
| 44 | Loire-Atlantique | Pays de la Loire | 1614 | 66 | 3.6% | [2.3-4.8] |
| 45 | Loiret | Centre-Val de Loire | 635 | 36 | 6.1% | [3.8-8.4] |
| 46 | Lot | Occitanie | 335 | 12 | 5.2% | [1-9.5] |
| 47 | Lot-et-Garonne | Nouvelle-Aquitaine | 279 | 12 | 2.4% | [0.9-3.9] |
| 48 | Lozère | Occitanie | 306 | 14 | 2.9% | [1.2-4.6] |
| 49 | Maine-et-Loire | Pays de la Loire | 788 | 35 | 3.5% | [2.1-4.8] |
| 50 | Manche | Normandie | 427 | 15 | 2.3% | [0.3-4.4] |
| 51 | Marne | Grand Est | 513 | 31 | 5.8% | [3.6-8] |
| 52 | Haute-Marne | Grand Est | 281 | 19 | 6.8% | [2.1-11.5] |

| | | | | | | |
|---|---|---|---|---|---|---|
| 53 | Mayenne | Pays de la Loire | 311 | 18 | 3.9% | [1.1-6.6] |
| 54 | Meurthe-et-Moselle | Grand Est | 664 | 33 | 3.6% | [1.7-5.5] |
| 55 | Meuse | Grand Est | 315 | 20 | 4.8% | [1.6-8.1] |
| 56 | Morbihan | Bretagne | 828 | 31 | 3.5% | [1.5-5.6] |
| 57 | Moselle | Grand Est | 930 | 60 | 7.3% | [5.5-9.1] |
| 58 | Nièvre | Bourgogne-Franche-Comté | 295 | 18 | 7.6% | [3.3-12] |
| 59 | Nord | Hauts-de-France | 2207 | 185 | 7.9% | [6.2-9.5] |
| 60 | Oise | Hauts-de-France | 1678 | 107 | 6.4% | [4.7-8.1] |
| 61 | Orne | Normandie | 241 | 10 | 3.0% | [0.9-5.1] |
| 62 | Pas-de-Calais | Hauts-de-France | 1140 | 71 | 4.9% | [3.3-6.6] |
| 63 | Puy-de-Dôme | Auvergne-Rhône-Alpes | 623 | 30 | 4.9% | [2.6-7.1] |
| 64 | Pyrénées-Atlantiques | Nouvelle-Aquitaine | 615 | 30 | 4.7% | [2.4-7.1] |
| 65 | Hautes-Pyrénées | Occitanie | 317 | 10 | 4.4% | [1.1-7.7] |
| 66 | Pyrénées-Orientales | Occitanie | 365 | 17 | 5.0% | [2.6-7.5] |
| 67 | Bas-Rhin | Grand Est | 1201 | 101 | 8.0% | [5.8-10.2] |
| 68 | Haut-Rhin | Grand Est | 1607 | 181 | 9.2% | [7.2-11.2] |
| 69 | Rhône | Auvergne-Rhône-Alpes | 1963 | 168 | 10.3% | [8.1-12.5] |
| 70 | Haute-Saône | Bourgogne-Franche-Comté | 304 | 22 | 4.6% | [2.5-6.7] |
| 71 | Saône-et-Loire | Bourgogne-Franche-Comté | 467 | 29 | 6.3% | [3.7-8.8] |
| 72 | Sarthe | Pays de la Loire | 458 | 15 | 1.6% | [0.3-3] |
| 73 | Savoie | Auvergne-Rhône-Alpes | 440 | 41 | 10.1% | [5.9-14.4] |
| 74 | Haute-Savoie | Auvergne-Rhône-Alpes | 754 | 72 | 10.0% | [7.8-12.2] |
| 75 | Paris | Ile-de-France | 2339 | 249 | 11.6% | [9.7-13.4] |
| 76 | Seine-Maritime | Normandie | 1032 | 40 | 3.1% | [1.7-4.6] |
| 77 | Seine-et-Marne | Ile-de-France | 1113 | 98 | 8.0% | [5.7-10.2] |
| 78 | Yvelines | Ile-de-France | 1418 | 112 | 9.5% | [7.4-11.7] |
| 79 | Deux-Sèvres | Nouvelle-Aquitaine | 329 | 11 | 2.7% | [0.6-4.8] |
| 80 | Somme | Hauts-de-France | 449 | 33 | 9.3% | [5.6-13.1] |
| 81 | Tarn | Occitanie | 301 | 13 | 2.7% | [0.9-4.4] |
| 82 | Tarn-et-Garonne | Occitanie | 251 | 6 | 1.6% | [0.2-2.9] |
| 83 | Var | Provence-Alpes-Côte d'Azur | 748 | 28 | 3.9% | [2.3-5.4] |
| 84 | Vaucluse | Provence-Alpes-Côte d'Azur | 422 | 16 | 2.4% | [0.4-4.3] |
| 85 | Vandée | Pays de la Loire | 698 | 14 | 2.2% | [0.6-3.7] |
| 86 | Vienne | Nouvelle-Aquitaine | 414 | 9 | 1.5% | [0.2-2.7] |
| 87 | Haute-Vienne | Nouvelle-Aquitaine | 315 | 9 | 2.9% | [0.7-5.1] |
| 88 | Vosges | Grand Est | 323 | 27 | 6.7% | [4.1-9.3] |
| 89 | Yonne | Bourgogne-Franche-Comté | 282 | 11 | 2.5% | [0.8-4.2] |
| 90 | Territoire de Belfort | Bourgogne-Franche-Comté | 316 | 26 | 7.1% | [4.5-9.7] |
| 91 | Essonne | Ile-de-France | 1118 | 99 | 12.6% | [9.5-15.7] |
| 92 | Hauts-de-Seine | Ile-de-France | 1589 | 138 | 9.0% | [6.8-11.2] |
| 93 | Seine-St-Denis | Ile-de-France | 850 | 108 | 12.5% | [9.6-15.4] |
| 94 | Val-de-Marne | Ile-de-France | 1178 | 124 | 12.7% | [10.1-15.4] |
| 95 | Val-D'Oise | Ile-de-France | 836 | 93 | 12.2% | [8.4-16] |
| 2A | Corse-du-Sud | Corse | 147 | 5 | 5.0% | [1.7-8.4] |
| 2B | Haute-Corse | Corse | 142 | 4 | 4.6% | [1.4-7.8] |

**Legend of Supplementary Table 1**

1. Home sampling by finger prick/Euroimmun ELISA-S test
2. People aged 15 years or over residing in mainland France, outside nursing homes and prisons.
3. The percentages are weighted by sampling weight (the reverse of inclusion probability), corrected for non-response probability and calibrated on the margin of the census. The prevalences are not equal to n/N.

**Supplementary Table 2: Factors associated with seropositivity (ELISA-S ≥ 1.1[1]) in November 2020 among people living in mainland France [2]. The national EpiCov cohort, 2020 November round – Univariate and multivariate analysis including detailed occupation, detailed living conditions and self-reported distancing behaviours over the last 7 days**

| | *Univariate analysis (proportion of Elisa-S ≥ 1.1)* | | | | | | *Logistic regressions[4]* | | | | |
|---|---|---|---|---|---|---|---|---|---|---|---|
| | N | n | % [3] | 95% CI[3] | *Crude OR* [3] | | *Model 1* | *Model 2* | *Model 3* | *Model 4* | *Model 5* |
| Immigration status | | | | | | | | | | | |
| No (majority population) | 54,296 | 3172 | 5.3 | [5.0-5.6] | ref | <0.001 | ref | ref | ref | ref | ref |
| First- generation from Europe | 1577 | 84 | 5.2 | [3.9-6.8] | 1.0 (0.7-1.3) | | 1.0 (0.7-1.3) | 0.9 (0.7-1.3) | 0.9 (0.7-1.3) | 0.9 (0.7-1.3) | 1.0 (0.7-1.3) |
| Second- generation Europe | 3164 | 197 | 6.0 | [4.9-7.3] | 1.1 (0.9-1.4) | | 1.1 (0.9-1.4) | 1.2 (0.9-1.5) | 1.2 (0.9-1.5) | 1.2 (0.9-1.5) | 1.2 (0.9-1.5) |
| First-generation from outside Europe | 1760 | 207 | 13.3 | [10.7-16.3] | 2.7 (2.1-3.5) | | 1.8 (1.4-2.4) | 1.6 (1.2-2.1) | 1.7 (1.3-2.2) | 1.6 (1.2-2.1) | 1.4 (1.1-1.9) |
| Second- generation outside Europe | 1894 | 233 | 14.4 | [11.9-17.4] | 3.0 (2.4-3.8) | | 1.9 (1.5-2.4) | 1.9 (1.5-2.4) | 1.9 (1.5-2.5) | 1.9 (1.5-2.5) | 2.0 (1.5-2.5) |
| *Demographic and socio-economic factors* | | | | | | | | | | | |
| Gender | | | | | | | | | | | |
| Men | 27564 | 1665 | 6.1 | [5.7-6.6] | ref | 0.46 | ref | ref | ref | ref | ref |
| Women | 35960 | 2278 | 6.4 | [6.0-6.8] | 1.0 (0.9-1.2) | | 1.1 (1.0-1.2) | 1.0 (0.9-1.1) | 1.0 (0.9-1.1) | 1.0 (0.9-1.1) | 1.0 (0.9-1.1) |
| Age in November | | | | | | | | | | | |
| 15-17 | 1438 | 128 | 9.8 | [7.8-12.2] | 2.8 (2.0-4.0) | <0.001 | 1.9 (1.3-2.8) | 2.1 (1.4-3.2) | 1.8 (1.2-2.8) | 2.0 (1.3-3.0) | 2.4 (1.6-3.6) |
| 18-24 | 4919 | 483 | 10.0 | [8.6-11.5] | 2.9 (2.2-3.9) | | 2.0 (1.5-2.7) | 2.0 (1.5-2.8) | 1.9 (1.4-2.7) | 2.0 (1.4-2.7) | 2.2 (1.6-3.0) |
| 25-34 | 6816 | 490 | 7.2 | [6.3-8.3] | 2.0 (1.5-2.7) | | 1.3 (0.9-1.8) | 1.4 (1.0-2.0) | 1.4 (1.0-2.0) | 1.5 (1.1-2.1) | 1.5 (1.0-2.1) |
| 35-44 | 10345 | 671 | 6.5 | [5.7-7.2] | 1.8 (1.4-2.4) | | 1.1 (0.8-1.5) | 1.2 (0.9-1.7) | 1.2 (0.9-1.7) | 1.3 (0.9-1.9) | 1.4 (1.0-2.0) |
| 45-54 | 12596 | 850 | 6.5 | [5.9-7.3] | 1.8 (1.4-2.4) | | 1.2 (0.9-8.1) | 1.4 (1.0-1.9) | 1.3 (0.9-1.8) | 1.4 (1.0-1.9) | 1.5 (1.0-2.0) |
| 55-64 | 12879 | 719 | 5.3 | [4.5-5.6] | 1.5 (1.1-1.9) | | 1.1 (0.9-1.5) | 1.2 (0.9-1.7) | 1.2 (0.9-1.6) | 1.2 (0.9-1.7) | 1.4 (1.0-1.8) |
| 65-74 | 10616 | 462 | 4.3 | [3.8-4.9] | 1.2 (0.9-1.6) | | 1.1 (0.9-1.5) | 1.1 (0.9-1.5) | 1.1 (0.8-1.5) | 1.2 (0.9-1.6) | 1.2 (0.9-1.6) |
| 75+ | 3920 | 149 | 3.7 | [2.9-4.7] | ref | | ref | ref | ref | ref | ref |
| Detailed occupation in November | | | | | | | | | | | |
| Physician (hospital) | 198 | 28 | 15.0 | [10.2-22.1] | 2.6 (1.6-4.1) | <0.001 | 2.5 (1.5-4.2) | 2.4 (1.4-4.0) | 2.3 (1.3-4.0) | 2.3 (1.3-4.0) | 2.2 (1.3-3.8) |
| Student (hospital) | 66 | 11 | 14.2 | [7.3-25.9] | 2.4 (1.1-5.1) | | 1.7 (0.7-4.1) | 1.7 (0.6-4.6) | 1.2 (0.3-3.9) | 1.2 (0.4-3.8) | 1.8 (0.7-4.7) |
| Nurse chief (hospital) | 147 | 25 | 17.1 | [9.4-29.2] | 3.0 (1.5-6.0) | | 3.3 (1.8-6.0) | 3.1 (1.8-5.4) | 3.5 (1.9-6.4) | 3.5 (1.9-6.4) | 3.1 (1.7-5.4) |
| Nurse (hospital) | 899 | 87 | 11.4 | [8.8-14.7] | 1.9 (1.4-2.5) | | 2.2 (1.6-2.9) | 1.8 (1.3-2.5) | 1.6 (1.1-2.3) | 1.6 (1.1-2.3) | 1.7 (1.2-2.4) |
| Assistant nurse (hospital) | 516 | 77 | 15.1 | [11.1-20.2] | 2.6 (1.8-3.7) | | 3.0 (2.1-4.4) | 3.0 (2.0-4.5) | 2.8 (1.7-4.5) | 2.8 (1.7-4.6) | 3.1 (2.0-4.7) |
| Other hospital personal | 260 | 14 | 6.5 | [3.2-12.6] | 1.0 (0.5-2.1) | | 1.3 (0.7-2.7) | 0.8 (0.4-1.6) | 0.8 (0.4-1.6) | 0.8 (0.4-1.6) | 0.7 (0.4-1.6) |
| Physician (non hosp) | 125 | 10 | 6.2 | [3.9-12.4] | 1.0 (0.5-2.1) | | 1.2 (0.5-2.5) | 1.1 (0.5-2.3) | 1.0 (0.5-2.4) | 1.1 (0.5-2.4) | 1.1 (0.5-2.3) |
| Nurse (private, non hosp) | 121 | 10 | 10.7 | [5.0-21.4] | 1.7 (0.8-3.9) | | 2.5 (1.1-5.8) | 2.5 (1.1-5.7) | 1.9 (0.9-4.0) | 1.9 (0.9-4.0) | 2.5 (1.1-5.7) |
| Pharmacist | 291 | 20 | 5.8 | [3.5-9.3] | 0.9 (0.5-1.5) | | 0.9 (0.5-1.6) | 0.8 (0.4-1.6) | 0.9 (0.5-1.6) | 0.9 (0.5-1.6) | 0.8 (0.4-1.6) |
| Teacher | 2343 | 136 | 5.9 | [4.9-7.0] | 1.0 (0.8-1.3) | | 1.0 (0.8-1.3) | 0.9 (0.7-1.2) | 0.9 (0.7-1.1) | 0.9 (0.7-1.1) | 1.0 (0.8-1.2) |
| Working in essential store | 402 | 24 | 4.9 | [3.0-7.7] | 0.8 (0.5-1.4) | | 0.8 (0.5-1.4) | 0.8 (0.5-1.4) | 0.9 (0.5-1.5) | 0.9 (0.5-1.5) | 0.9 (0.5-1.5) |
| Other essential | 630 | 52 | 7.5 | [5.1-11.1] | 1.1 (0.7-1.8) | | 1.1 (0.7-1.8) | 1.2 (0.8-2.0) | 1.3 (0.8-2.2) | 1.3 (0.8-2.3) | 1.3 (0.8-2.1) |
| Unclear, not recoded | 1751 | 122 | 8.2 | [6.1-10.9] | 1.3 (0.9-1.8) | | 1.3 (0.9-1.8) | 1.3 (1.0-1.8) | 1.4 (1.0-2.0) | 1.4 (1.0-2.0) | 1.3 (1.0-1.1) |
| Non-essential | 24092 | 1685 | 6.5 | [6.0-7.0] | ref | | ref | ref | ref | ref | ref |
| Not working | 28593 | 1535 | 5.6 | [5.2-6.0] | 0.9 (0.8-1.1) | | 0.9 (0.8-1.1) | 1.0 (0.8-1.1) | 1.0 (0.8-1.1) | 1.0 (0.8-1.1) | 0.9 (0.8-1.1) |
| Highest diploma attained | | | | | | | | | | | |
| No diploma | 2657 | 166 | 6.2 | [5.0-7.7] | ref | <0.001 | | ref | ref | | |
| Primary education | 5839 | 322 | 5.1 | [4.4-5.9] | 0.8 (0.6-1.1) | | | 0.9 (0.6-1.1) | 0.8 (0.6-1.1) | 0.8 (0.6-1.1) | 0.8 (0.6-1.1) |
| Vocational secondary | 11483 | 580 | 4.8 | [4.2-5.5] | 0.8 (0.6-1.0) | | | 0.9 (0.7-1.2) | 1.0 (0.7-1.3) | 1.0 (0.7-1.3) | 0.9 (0.7-1.2) |
| high school diploma | 12765 | 817 | 6.7 | [6.0-7.5] | 1.1 (0.8-1.4) | | | 1.0 (0.8-1.3) | 1.0 (0.8-1.4) | 1.0 (0.8-1.4) | 1.0 (0.8-1.3) |

| | | | | | | | | | | | |
|---|---|---|---|---|---|---|---|---|---|---|---|
| High school + 2 to 4 years | 19210 | 1245 | 7.5 | [6.9-8.1] | 1.2 (1.0-1.6) | | | 1.1 (0.8-1.5) | 1.2 (0.9-1.6) | 1.2 (0.9-1.6) | 1.1 (0.8-1.5) |
| High school + 5 years or more | 11570 | 813 | 7.0 | [6.4-7.6] | 1.1 (0.9-1.5) | | | 0.9 (0.7-1.2) | 1.0 (0.7-1.3) | 1.0 (0.7-1.3) | 0.9 (0.7-1.3) |
| Family income per capita (deciles) | | | | | | | | | | | |
| D01(lowest) | 3672 | 241 | 8.2 | [6.7-10.0] | 1.4 (1.1-1.8) | <0.001 | | 1.1 (0.9-1.4) | 1.1 (0.9-1.5) | 1.1 (0.9-1.4) | 1.1 (0.8-1.4) |
| D02-D03 | 6481 | 385 | 6.2 | [5.3-7.3] | 1.1 (0.9-1.3) | | | 0.9 (0.7-1.1) | 0.9 (0.7-1.1) | 0.9 (0.7-1.1) | 0.9 (0.7-1.0) |
| D04-D05 | 9098 | 523 | 5.8 | [4.4-6.5] | 0.9 (0.8-1.0) | | | 0.9 (0.7-1.0) | 0.8 (0.7-1.0) | 0.9 (0.7-1.0) | 0.9 (0.7-1.0) |
| D06-D07 | 13252 | 784 | 5.9 | [5.4-6.5] | ref | | | ref | ref | ref | ref |
| D08-D09 | 18724 | 1147 | 6.1 | [5.7-6.6] | 1.0 (0.9-1.2) | | | 1.0 (0.9-1.2) | 1.0 (0.9-1.1) | 1.0 (0.9-1.2) | 1.0 (0.9-1.1) |
| D10 | 10880 | 766 | 7.0 | [6.5-7.6] | 1.2 (1.1-1.4) | | | 1.1 (0.9-1.3) | 1.0 (0.9-1.2) | 1.0 (0.9-1.2) | 1.1 (0.9-1.2) |
| *Living conditions* | | | | | | | | | | | |
| Urban areas according to the proportions of immigrant populations | | | | | | | | | | | |
| Paris | 11122 | 1049 | 10.7 | [9.7-11.8] | 2.5 (2.2-2.8) | <0.001 | 1.1 (0.8-1.5) | 1.1 (0.8-1.5) | 1.1 (0.9-1.5) | 1.1 (0.9-1.5) | 1.1 (0.8-1.5) |
| Lyon | 2370 | 206 | 10.7 | [8.6-13.3] | 2.5 (1.9-3.2) | | 1.3 (1.0-1.7) | 1.3 (0.9-1.7) | 1.3 (1.0-1.8) | 1.3 (1.0--1.7) | 1.2 (0.9-1.6) |
| Marseille | 1345 | 83 | 6.5 | [4.6-9.1] | 1.4 (1.0-2.1) | | 1.2 (0.8-1.8) | 1.3 (0.9-1.9) | 1.2 (0.8-1.9) | 1.2 (0.8-1.8) | 1.2 (0.8-1.9) |
| Nice | 808 | 32 | 3.0 | [2.0-4.5] | 0.6 (0.4-1.0) | | 0.8 (0.5-1.2) | 0.8 (0.5-1.3) | 0.8 (0.5-1.3) | 0.8 (0.5-1.3) | 0.9 (0.6-1.3) |
| Toulouse | 1503 | 76 | 5.8 | [4.1-8.1] | 1.3 (0.9-1.8) | | 1.2 (0.9-1.7) | 1.0 (0.8-1.4) | 1.1 (0.8-1.5) | 1.1 (0.8-1.5) | 1.0 (0.7-1.4) |
| Lille | 1098 | 102 | 9.7 | [7.1-13.0] | 2.2 (1.6-3.1) | | 1.3 (0.9-2.0) | 1.3 (0.8-1.9) | 1.2 (0.8-1.5) | 1.2 (0.8-1.9) | 1.3 (0.8-1.9) |
| Strasbourg | 835 | 74 | 8.8 | [6.6-11.8] | 2.0 (1.4-2.8) | | 1.2 (0.8-1.7) | 1.2 (0.8-1.7) | 1.2 (0.9-1.8) | 1.2 (0.9-1.8) | 1.2 (0.8-1.7) |
| Bordeaux | 1217 | 48 | 3.8 | [2.6-5.6] | 0.8 (0.5-1.2) | | 0.9 (0.6-1.4) | 0.9 (0.6-1.4) | 0.9 (0.6-1.4) | 0.9 (0.6-1.4) | 0.8 (0.5-1.2) |
| Grenoble | 795 | 53 | 8.3 | [5.4-11.4] | 1.8 (1.2-2.7) | | 1.0 (0.7-1.6) | 1.0 (0.7-1.6) | 1.0 (0.6-1.6) | 1.0 (0.6-1.6) | 1.0 (0.6-1.6) |
| Genève-Annemasse | 293 | 27 | 6.2 | [5.1-13.3] | 1.9 (1.1-3.2) | | 1.0 (0.6-1.8) | 1.0 (0.6-1.8) | 1.1 (0.6-2.1) | 1.1 (0.6-2.1) | 1.1 (0.6-2.0) |
| All other places in France | 42075 | 2187 | 4.7 | [4.4-5.0] | ref | | ref | ref | ref | ref | ref |
| Regions the most affected by Covid according to registered hospitalization rates from the start of the epidemic | | | | | | | | | | | |
| Regions with lowest incidence | 32472 | 1360 | 3.8 | [3.6-4.2] | ref | <0.001 | ref | ref | ref | ref | ref |
| Auvergne Rhone Alpes | 8274 | 643 | 8.4 | [7.4-9.4] | 2.3 (2.0-2.7) | | 2.0 (1.7-2.5) | 2.0 (1.7-2.5) | 1.9 (1.5-2.3) | 1.9 (1.5-2.3) | 2.1 (1.7-2.6) |
| Hauts-de-France | 5876 | 418 | 6.8 | [5.9-7.9] | 1.8 (1.5-2.2) | | 1.6 (1.3-2.0) | 1.6 (1.3-2.0) | 1.5 (1.2-1.9) | 1.5 (1.2-1.9) | 1.7 (1.3-2.1) |
| Ile-de-France | 10441 | 1021 | 11.0 | [10.0-12.2] | 3.1 (2.7-3.6) | | 2.2 (1.6-2.9) | 2.1 (1.6-3.0) | 1.9 (1.4-2.6) | 1.9 (1.4-2.6) | 2.1 (1.5-2.9) |
| Grand Est | 6461 | 501 | 6.7 | [5.9-7.6] | 1.8 (1.5-2.1) | | 1.8 (1.5-2.1) | 1.8 (1.5-2.1) | 1.7 (1.4-2.1) | 1.8 (1.4-2.1) | 1.8 (1.5-2.2) |
| Population density in municipality | | | | | | | | | | | |
| Low | 23647 | 1178 | 4.5 | [4.1-4.8] | ref | <0.001 | ref | ref | ref | ref | ref |
| Medium | 18650 | 1075 | 5.4 | [4.9-6.0] | 1.2 (1.1-1.4) | | 1.1 (0.9-1.2) | 1.0 (0.9-1.2) | 1.0 (0.9-1.2) | 1.0 (0.9-1.2) | 1.1 (0.9-1.2) |
| High | 21227 | 1690 | 8.5 | [7.9-9.2] | 2.0 (1.8-2.2) | | 1.2 (1.0-1.4) | 1.2 (1.0-1.4) | 1.1 (1.0-1.3) | 1.1 (1.0-1.3) | 1.2 (1.1-1.4) |
| Socially deprived neighbourhood | | | | | | | | | | | |
| No | 61840 | 3778 | 5.9 | [5.6-6.2] | ref | <0.001 | ref | ref | ref | ref | ref |
| Yes | 1684 | 165 | 11.0 | [8.9-14.0] | 2.0 (1.5-2.6) | | 1.3 (1.0-1.7) | 1.4 (1.1-1.9) | 1.4 (1.1-1.9) | 1.5 (1.1-2.0) | 1.3 (1.0-1.8) |
| Number of people in household | | | | | | | | | | | |
| 1 | 10377 | 570 | 5.1 | [4.5-5.9] | ref | <0.001 | ref | ref | ref | ref | ref |
| 2 | 24994 | 1331 | 4.9 | [4.6-5.3] | 1.0 (0.8-1.1) | | 1.0 (0.9-1.2) | 1.0 (0.9-1.2) | 0.8 (0.7-0.9) | 0.8 (0.7-0.9) | 1.0 (0.9-1.2) |
| 3 | 10902 | 741 | 6.5 | [5.8-7.2] | 1.3 (1.1-1.5) | | 1.2 (1.0-1.4) | 1.1 (0.9-1.3) | 0.8 (0.7-1.0) | 0.8 (0.7-1.0) | 1.1 (0.9-1.4) |
| 4 | 12040 | 899 | 7.9 | [7.0-8.7] | 1.6 (1.3-1.9) | | 1.3 (1.1-1.6) | 1.3 (1.1-1.6) | 0.9 (0.7-1.1) | 0.9 (0.7-1.1) | 1.3 (1.1-1.5) |
| 5 or more | 5189 | 400 | 10 | [8.7-11.8] | 2.1 (1.7-2.6) | | 1.4 (1.1-1.8) | 1.3 (1.1-1.7) | 0.9 (0.7-1.1) | 0.9 (0.7-1.1) | 1.4 (1.1-1.7) |
| ≥ 1 household member tested positive | | | | | | | | | | | |
| No | 59740 | 2860 | 4.9 | [4.6-5.2] | ref | <0.001 | | | ref | ref | |
| Yes | 3724 | 1076 | 29.0 | [26.8-31.3] | 7.9 (7.0-9.0) | | | | 7.3 (6.3-8.4) | 7.2 (6.2-8.3) | |

| | | | | | | | | | | |
|---|---|---|---|---|---|---|---|---|---|---|
| *Individual factors* | | | | | | | | | | |
| Tobacco use | | | | | | | | | | |
| Daily smoker | 9056 | 255 | 2.6 | [2.1-3.0] | ref | <0.001 | ref | ref | ref | ref |
| Occasional smoker | 2322 | 146 | 6.8 | [5.0-8.6] | 3.1 (2.3-4.2) | | 2.5 (1.8-3.5) | 2.7 (1.9-3.7) | 2.6 (1.9-3.7) | 2.6 (1.8-3.5) |
| Ex smoker | 21767 | 1370 | 6.1 | [5.6-6.6] | 2.2 (1.8-2.7) | | 2.5 (2.0-3.1) | 2.5 (2.0-3.2) | 2.5 (2.0-3.2) | 2.4 (1.9-3.0) |
| Non-smoker | 30237 | 2163 | 7.6 | [7.1-8.1] | 3.0 (2.5-3.6) | | 2.9 (2.4-3.5) | 3.0 (2.4-3.7) | 3.0 (2.4-3.7) | 2.7 (2.2-3.3) |
| Body mass index | | | | | | | | | | |
| Low | 2187 | 135 | 6.2 | [4.9-8.0] | 1.0 (0.8-1.4) | 0.74 | 1.0 (0.8-1.4) | 1.1 (0.8-1.4) | 1.1 (0.8-1.4) | 1.1 (0.8-1.4) |
| Normal | 33232 | 2090 | 6.4 | [6.0-6.8] | ref | | ref | ref | ref | ref |
| High | 19123 | 1156 | 6.0 | [5.4-6.6] | 1.0 (0.7-1.3) | | 1.1 (0.8-1.5) | 1.2 (0.8-1.5) | 1.1 (0.8-1.5) | 1.1 (0.8-1.5) |
| Very high | 8748 | 545 | 6.2 | [5.9-6.6] | 1.0 (0.8-1.4) | | 1.2 (0.8-1.6) | 1.2 (0.8-1.6) | 1.2 (0.8-1.6) | 1.1 (0.8-1.6) |
| History of diabetes | | | | | | | | | | |
| No | 60690 | 3793 | 6.3 | [6.30-6.6] | ref | 0.18 | ref | ref | ref | |
| Yes | 2803 | 2090 | 5.3 | [4.1-6.8] | 0.8 (0.6-1.1) | | 0.9 (0.7-1.3) | 0.9 (0.7-1.2) | 0.9 (0.6-1.2) | |
| In the previous 7 days: | | | | | | | | | | |
| Outing in street without mask | | | | | | | | | | |
| No outing | 1993 | 244 | 9.2 | [7.6-11.0] | 1.4 (1.2-1.8) | <0.001 | | | 1.5 (1.2-1.9) | 1.6 (1.2-2.0) |
| Systematic mask use | 40234 | 2557 | 6.6 | [6.2-7.0] | ref | | | | ref | ref |
| No or non-systematic use | 21297 | 1142 | 5.0 | [4.6-5.4] | 0.7 (0.7-0.8) | | | | 0.9 (0.8-1.0) | 0.9 (0.7-1.1) |
| Familial outing without mask | | | | | | | | | | |
| No outing | 36950 | 2481 | 6.8 | [6.4-7.2] | 1.2 (1.0-1.4) | <0.001 | | | 1.1 (0.9-1.3) | 1.0 (0.8-1.3) |
| Systematic mask use | 6128 | 341 | 5.9 | [5.0-7.0] | ref | | | | ref | ref |
| No or non-systematic use | 20446 | 1121 | 5.2 | [4.7-5.6] | 0.9 (0.7-1.1) | | | | 1.0 (0.8-1.2) | 0.9 (0.7-1.1) |
| Festive outing without mask | | | | | | | | | | |
| No outing | 60246 | 3698 | 6.2 | [5.9-6.6] | 1.0 (0.6-1.7) | 0.13 | | | 0.9 (0.5-1.5) | 0.9 (0.6-1.6) |
| Systematic mask use | 383 | 27 | 6.1 | [3.8-9.4] | ref | | | | ref | ref |
| No or non-systematic use | 2895 | 218 | 7.4 | [6.2-8.8] | 1.2 (0.7-2.1) | | | | 1.1 (0.7-1.9) | 1.3 (0.8-2.2) |
| Number of prolonged contacts | | | | | | | | | | |
| No contact | 7964 | 538 | 6.3 | [5.5-7.2] | 0.9 (0.7-1.1) | 0.10 | | | | 0.8 (0.7-1.0) |
| No prolonged contacts | 13801 | 781 | 5.6 | [5.0-6.2] | ref | | | | | ref |
| < 5 | 34578 | 2147 | 6.3 | [5.9-6.7] | 1.0 (0.9-1.2) | | | | | 0.9 (0.7-1.0) |
| 5 to <10 | 4222 | 279 | 7.4 | [6.0-9.1] | 1.2 (0.9-1.6) | | | | | 0.8 (0.6-1.0) |
| 10 to < 50 | 700 | 45 | 6.7 | [4.5-10.0] | 1.1 (0.7-1.7) | | | | | 0.7 (0.4-1.1) |
| 50 to < 100 | 51 | 1 | 1.5 | [0.2-9.9] | 0.2 (0.03-1.6) | | | | | 0.2 (0.03-1.5) |
| ≥ 100 | 28 | 2 | 6.2 | [1.6-24.0] | 1.1 (0.2-4.7) | | | | | 0.8 (0.2-4.3) |

**Legend of Supplementary Table 2**

1. Home sampling by finger prick/Euroimmun ELISA-S test
2. People aged 15 years or over residing in mainland France, outside nursing homes and prisons.
3. The sampling design is taken into account for the estimation of prevalence, confidence intervals (logit transformation), crude and adjusted odds ratios, confidence intervals and tests, statistical tests, with the SAS procsurvey procedure. The percentages are weighted by sampling weight (the reverse of inclusion probability), corrected for non-response probability and calibrated on the margin of the census. The prevalences are not equal to n/N.

4. MODELS : All models were adjusted for immigration status, gender, age, detailed occupation, urban areas with the most immigration, regions with highest hospitalization rates, population density in the municipality, socially-deprived neighbourhood and household size.

   Model 2 additionally adjusted for highest diploma attained, family income per capita, tobacco use and body mass index.
   Model 3 = model 2 + presence of one positive member in the household,
   Model 4 = model 3 + self-reported distancing behaviours over the last 7 days: outing in street without mask, familial outing without mask, festive outing without mask
   Model 5 = model 3 + festive outing without mask and number of prolonged contacts, and without presence of one positive household member
5. Migratory status: Majority population = persons born in France who are neither first nor second-generation immigrants / First-generation immigrants: born non-French outside France and living permanently in France (including those who subsequently acquired French nationality) / Second-generation immigrants: born and living in France, with at least one parent being a first-generation immigrant
